# A Genome-Wide Association Study of Survival in Patients with Sepsis

**DOI:** 10.1101/2022.05.06.22274756

**Authors:** Tamara Hernandez-Beeftink, Beatriz Guillen-Guio, Jose M. Lorenzo-Salazar, Almudena Corrales, Eva Suarez-Pajes, Rui Feng, Luis A. Rubio-Rodríguez, Megan L Paynton, Raquel Cruz-Guerrero, M. Isabel García-Laorden, Miryam Prieto-González, Aurelio Rodríguez-Pérez, Demetrio Carriedo, Jesús Blanco, Alfonso Ambrós, Elena González-Higueras, Elena Espinosa, Arturo Muriel, Eduardo Tamayo, María M Martin, Leonardo Lorente, David Domínguez, Abelardo García de Lorenzo, Heather M. Giannini, John P. Reilly, Tiffanie K. Jones, José M. Añón, Marina Soro, Ángel Carracedo, Louise V. Wain, Nuala J Meyer, Jesús Villar, Carlos Flores, the Genetics of Sepsis (GEN-SEP) Network

**Author notes:** **Correspondence:** Dr. Carlos Flores, Research Unit, Hospital Universitario Nuestra Señora de Candelaria, Carretera del Rosario s/n, 38010 Santa Cruz de Tenerife, Spain.

## Abstract

**Background:** Sepsis is a severe systemic inflammatory response to infections that is accompanied by organ dysfunction and has a high mortality rate in adult intensive care units (ICUs). Most genetic studies have identified gene variants associated with development and outcomes of sepsis focusing on biological candidates. We conducted the first genome-wide association study (GWAS) of 28-day survival in adult patients with sepsis.

**Methods:** This study was conducted in two stages. The first stage was performed on 687 European sepsis patients from the GEN-SEP network and 7.5 million imputed variants. Association testing was conducted with Cox regression models, adjusting by sex, age, and the main principal components of genetic variation. A second stage focusing on the prioritized genetic variants was performed on 2,063 ICU sepsis patients (1,362 European Americans and 701 African Americans) from the MESSI study. A meta-analysis of results from the two stages was conducted and significance was established at *p*<5.0×10^−8^. Whole-blood transcriptomic and functional annotations were evaluated on the identified genes and variants.

**Findings:** We identified three independent variants associated with reduced 28-day sepsis survival, including a missense variant in *SAMD9* (hazard ratio [95% confidence interval]=1.64 [1.37-6.78], *p*=4.92×10^−8^). *SAMD9* encodes a mediator of the inflammatory response to tissue injury that is overexpressed in peripheral blood of non-surviving sepsis patients compared to those surviving (*p*=2.18×10^−3^).

**Interpretation:** We performed the first GWAS of 28-day sepsis survival and identified novel variants associated with reduced survival. Our findings could allow the identification of novel targets for sepsis treatment and patient risk stratification.

**Research in context:** *Evidence before this study:* Sepsis is defined as a life-threatening clinical syndrome of physiological, pathological, and biochemical abnormalities caused by a dysregulated host response to an infection, and with long-term physical, psychological, and cognitive disabilities. Many genetic studies have focused on identifying genetic risk factors associated with sepsis development and severity, but only four genome-wide association studies (GWAS) have been published to date. Three of them focused on sepsis mortality. The first study identified that common genetic variation in the *FER* gene associated with a reduced risk of death. The second study found variants associated with an increased risk of death in *VPS13A*, which is key in autophagic degradation. In the last study, variants of the *CISH* gene, involved in cytokine regulation, were associated with the risk of death. Nevertheless, there is a lack of GWAS focused on sepsis survival, which takes into account the probability estimates of death for each patient over time.

*Added value of this study:* To the best of our knowledge, we provide the results of the first GWAS of 28-day sepsis survival conducted to date. In this two-staged study, we identified three novel loci associated with reduced 28-day survival among sepsis patients. We identified one missense variant in *SAMD9*, which encodes a critical regulator in the inflammatory response and apoptosis. A significant upregulation of *SAMD9* gene expression in whole blood was observed among non-surviving sepsis patients compared to those surviving. Associations were also found for one intergenic variant to *SLC5A12\FIBIN* and an intergenic variant to two non-coding RNAs (LINC00378\MIR3169).

*Implications of all the available evidence:* The identification of effective prognostic genetic markers in sepsis is a promising instrument for clinical practice. This study identified three novel genetic factors of fatal outcomes, all having interesting and important biological plausibly that could serve as novel targets for sepsis treatment. This knowledge is important to propose effective sepsis treatments and will be central in the development of personalized medicine approaches.

## Introduction

Sepsis is defined as a severe systemic inflammatory response to infections that is accompanied by organ dysfunction^1^. It is recognized as a global priority and its incidence in adults is estimated at approximately 189 cases per 100,000 people per year^2-3^. In the intensive care units (ICUs), sepsis is associated with an overall mortality rate of about 30%^4-5^ and with significant morbidity in survivors. The risk of death in patients with sepsis increases with hemodynamic instability (i.e. septic shock) or due to respiratory complications, such as the acute respiratory distress syndrome (ARDS)^6^. Given the lack of specific therapeutic options and the underlying etiological complexity, multiple studies have focused on improving prevention, diagnosis, and prognosis of sepsis^5^.

Genetic variation influences the host immune response to microbial agents^7-10^. In this sense the genome-wide association studies (GWAS) have an enormous potential to reveal genetic factors for disease susceptibility, severity, and/or survival, as it has been shown for a number of infectious diseases^10-14^. However, the number of GWAS of sepsis or its complications is limited. To date, only three GWAS have been completed for sepsis mortality, although the likelihood of death for each patient over time was not considered^15-17^. Specifically, Rautanen et al^15^ analysed 28-day mortality in patients with pneumonia, linking the FER tyrosine kinase (*FER*) gene variation with reduced risk for death from sepsis. Nevertheless, another study was unable to replicate this finding in independent patients^18^. Additionally, Scherag and colleagues found that the top ranking variants associated with 28-day mortality from sepsis in their study were located in the Vacuolar Protein Sorting 13 Homolog A (*VPS13A*) gene^16^. Finally, Rosier and colleagues identified variants within the Cytokine Inducible SH2 Containing Protein (*CISH*) associated with mortality due to septic shock at day 7 or day 28^17^.

Based on all the above-mentioned evidence, here we performed a GWAS of 28-day sepsis survival to identify novel genetic variants associated with sepsis outcome taking into account the probability estimates of death for each patient over time.

## Methods

### Study design and participants

We performed a GWAS of 28-day survival in patients with sepsis. This study was conducted in two stages. The first stage was based on two cohorts of patients from the GEN-SEP study^19^, where association results were meta-analysed and used to prioritize variants (**Figure 1**). In the second stage, we followed up these variants in independent sepsis patients of European and African American ancestry from the MESSI study^20^. Finally, a meta-analysis of results of the 2,750 patients (1,121 non-survivors) from the two stages was done and genome-wide significance was established at *p*<5.0×10^−8^.

**Figure 1.**
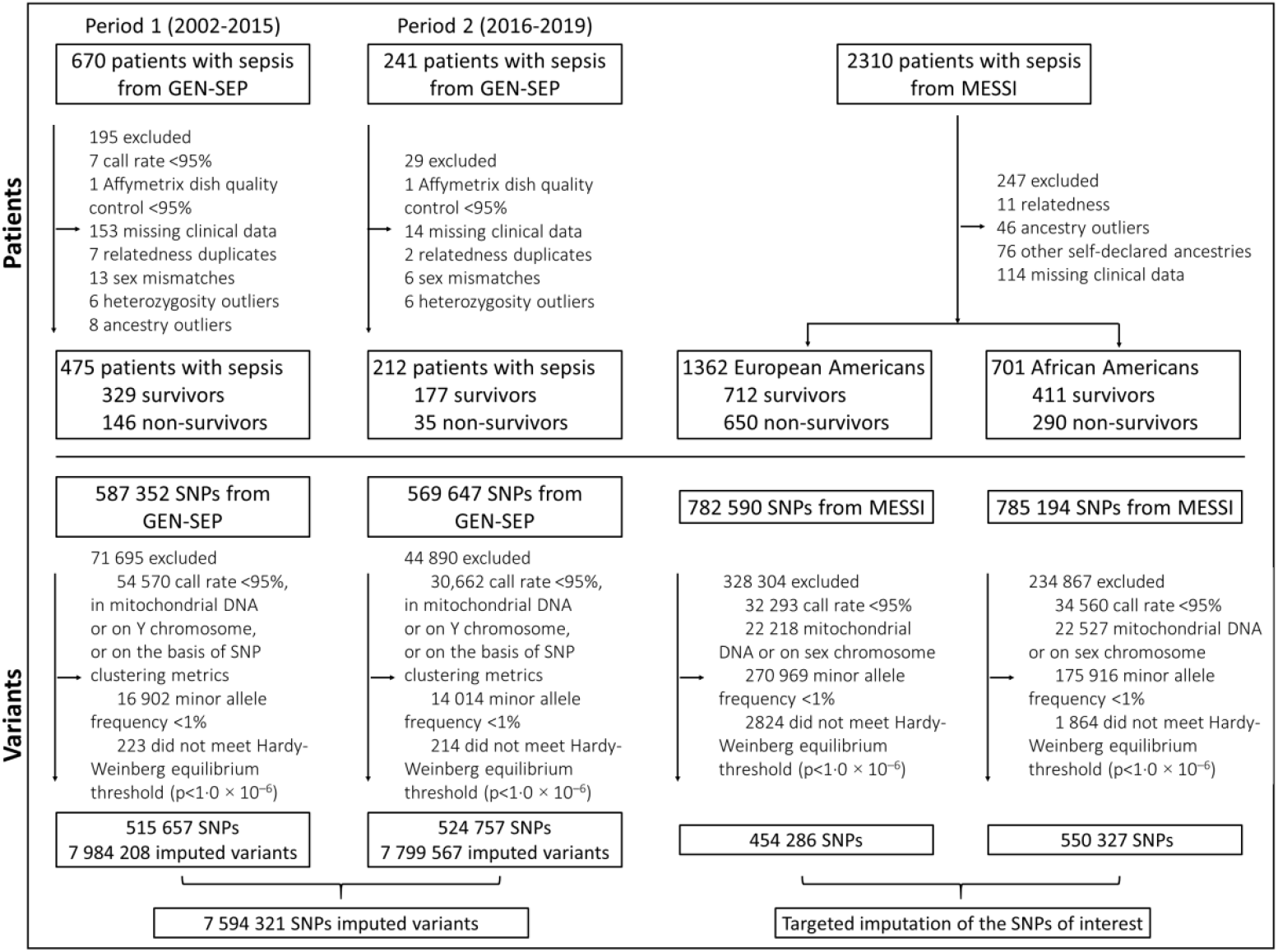
Study profile. SNPs, single-nucleotide polymorphisms.

The GEN-SEP study, used for the first stage, included 475 (146 non-survivors) European ancestry patients considered for a previous GWAS of sepsis associated ARDS^19^ recruited between 2002 and 2015 for which follow up records of 28-days survival were available (1st GEN-SEP period). In a second round of patient recruitment, between 2016 and 2019, another 212 (35 non-survivors) patients were included in the GEN-SEP study (2nd GEN-SEP period) (**Figure 1**) (see supplementary material for further details). Sepsis diagnosis was clinically defined according to the Third International Consensus Definitions for Sepsis^1^.

The MESSI study^20^, used for the second stage, included a total of 1,362 (650 non-survivors) unrelated European Americans and 701 (290 non-survivors) African Americans with a period of recruitment from 2008 to 2019 (see supplementary material for further details).

All participating studies were done according to The Code of Ethics of the World Medical Association (Declaration of Helsinki), and written, informed consent was obtained from all participants or their representatives. The Research Ethics Committees at all participating centers approved this study.

### Genotyping and statistical analyses

Genotyping in the GEN-SEP was performed using the Axiom Genome-Wide Human CEU 1 array (Affymetrix, Santa Clara, CA, USA). Genotyping quality control procedures are detailed in the supplementary material (**Figure 1**). We also calculated the main axis of genetic variation using principal component (PC) analyses (**Figure S1**). In MESSI, SNPs were genotyped using the Affymetrix Axiom TxArray v.1 (Affymetrix) (see supplementary material for further details).

In the first stage, a Cox proportional hazards regression model was used to test genetic associations adjusting for age, sex, and the first two main PCs. Results were obtained for a total of 7,872,728 (1^st^ GEN-SEP period) and 7,829,916 (2^nd^ GEN-SEP period) SNPs. An intersection of 7,682,187 SNPs was considered for fixed-effect model meta-analysis. Variants were prioritized for the next stage if they satisfied having the same effect direction, a *p*<0.05 in both GEN-SEP recruitment periods, and a *p*<5.0×10^−7^ after the meta-analysis of both periods. Association results of this first stage were also inspected to evaluate whether the variants or genes previously associated with sepsis mortality by other studies were also associated in GEN-SEP^15-17^.

In the second stage, the prioritized independent variants were tested for association in the MESSI European Americans and African Americans, separately, using Cox regression models, also considering sex, age, and the first two main PCs as covariates.

Finally, a fixed-effect model meta-analysis from the GEN-SEP and MESSI studies was performed, and the genome-wide significance was declared at *p*<5.0×10^−8^. A sensitivity analysis was conducted for the genome-wide significant variants, adjusting the models for different clinical and demographic variables and the index event bias. More details are included in the supplementary material.

### Association studies in the HLA genes

Given the importance of the major histocompatibility complex (MHC) in inflammatory and immunological diseases, we performed association testing with 28-day sepsis survival of genetic variation in the HLA region. Association analyses were performed only on the GEN-SEP cohort by using Cox regressions adjusting for sex, age, and the main two PCs. This analysis was restricted to 207 classic HLA alleles and 1,034 amino acids that had a frequency >1%. Considering the multiple tests adjustment, significance thresholds for the HLA analysis were set at *p*<2.49×10^−4^ and *p*<4.83×10^−5^, respectively.

### Annotation of the functional effects of associated variants and gene expression

The functional effects of the independent most significant (sentinel) variants, their linkage disequilibrium proxies (r^2^>0.7), and related genes were assessed based on empirical data from different integrated software tools and datasets (see supplementary material for further details). To assess differential gene expression of the genes near the sentinel variants, we accessed the public gene expression datasets GSE54514 and GSE32707, containing data for sepsis survival and sepsis-associated ARDS, respectively.

### Polygenic risks of sepsis and effects on 28-day sepsis survival

We examined whether the polygenic component of sepsis risk was associated with 28-day sepsis survival through polygenic risk scores (PRS) (see the supplementary material for further details).

First, we obtained a model of the genetic risk for sepsis by a GWAS of all available sepsis cases from GEN-SEP and population controls (**Table S1**) using logistic regressions adjusted by sex, age, and the two main PCs.

Then, we constructed the PRS for sepsis risk by including in the score those variants that met a *p*-value threshold in the sepsis risk GWAS and varied this threshold to investigate the effect of including more variants in the score.

Finally, we tested if the score was associated with 28-day survival among GEN-SEP patients, adjusting for sex, age, and two main PCs. For this, we used Cox regression and established the *p*<0.001 threshold for defining significant associations of the risk score.

Additionally, we performed sensitivity analyses to assess the sepsis risk score association with 28-day sepsis survival after 1) excluding variants that deemed significantly associated with sepsis in candidate gene studies (**Table S2**), and 2) excluding variants significantly associated with sepsis mortality in previous GWAS (see the supplementary material for further details).

## Results

Demographic and clinical features of patients from the first stage are described in **Table S3**. After association testing, 11 independent variants were prioritized in the first stage (**Figure 2; Table S4**). The genomic inflation factor of the results from this stage (λ=1.06) did not indicate major systematic deviations from the null hypothesis of no association (**Figure S2**).

**Figure 2.**
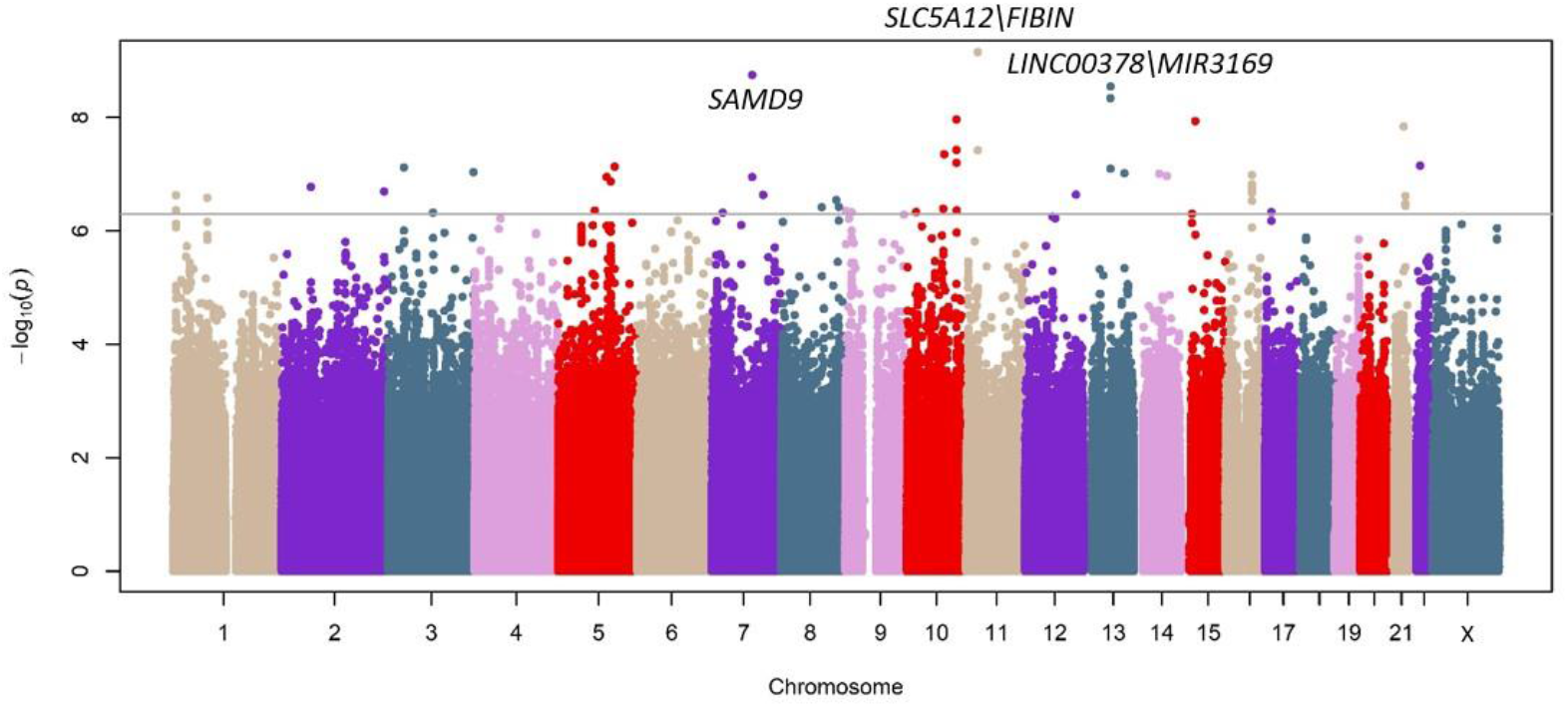
Association results for the 28-day sepsis survival for the first stage. Manhattan plot representing in the x-axis the genomic positions and in the y-axis the significance (-log10(p-value)). The horizontal line indicates the significance threshold for prioritization to the second stage (p=5.0×10^−7^).

In the second stage, we were able to follow up on the association of 10 of the 11 prioritized variants (**Table S4**). Three of these variants reached the genome-wide significance threshold in the meta-analysis (**Table 1**): a missense variant in the Sterile Alpha Motif Domain containing 9 (*SAMD9*) (rs34896991; p.Ala1556Thr), an intergenic variant to Solute Carrier family 5 member 12 (*SLC5A12*) and Fin Bud Initiation Factor Homolog (*FIBIN*) genes (rs146257041), and an intergenic variant between two non-coding RNAs (LINC00378 and MIR3169) (rs138347802). Regional association results and Kaplan-Meier survival plots for the three genome-wide significant variants in the GEN-SEP are reported in the supplementary material (**Figure S3**; **Figure S4**). Results remained robust after sensitivity analyses (**Table 2**; **Table S5**).

**Table 1.** Prioritized independent SNPs from the first stage of the GWAS of 28-day sepsis survival.

|  |  |  |  |  |  |  |  |  |  |  |  |  |  |
| --- | --- | --- | --- | --- | --- | --- | --- | --- | --- | --- | --- | --- | --- |
| rsID | Position (Hg19) | A1/A2 | Func.Gene | Gene | EAF | GEN-SEP (N=687) |  | MESSI (N=1,362)<br>(European Americans) |  | MESSI (N=701)<br>(African-Americans) |  | Meta-analysis (N=2,750) |  |
|  |  |  |  |  |  | HR (95% CI) | p-value | HR (95% CI) | P-value | HR (95% CI) | P-value | HR (95% CI) | p-value |
| rs34896991 | 7:92730745 | C/T | Exonic | *SAMD9* | 0.018 | 4.75 (2.86-7.89) | 1.77x10<sup>-9</sup> | 0.61 (0.29-1.28) | 0.188 | 1.44 (0.46-4.50) | 0.534 | 1.64 (1.37-6.78) | 4.92x10<sup>-8</sup> |
| rs146257041 | 11:26983813 | A/G | Intergenic | *SLC5A12\FIBIN* | 0.015 | 5.14 (3.06-8.65) | 7.04x10<sup>-10</sup> | NA | NA | 0.89 (0.12-6.39) | 0.911 | 4.59 (2.77-7.59) | 3.00x10<sup>-9*</sup> |
| rs138347802 | 13:61367197 | A/G | Intergenic | *LINC00378\MIR3169* | 0.014 | 8.33 (4.1-16.91) | 4.56x10<sup>-9</sup> | 0.68 (0.30-1.52) | 0.345 | 2.98 (0.95-9.38) | 0.062 | 2.57 (1.82-13.03) | 4.44x10<sup>-8</sup> |
| *Meta-analysis performed between two studies. |  |  |  |  |  |  |  |  |  |  |  |  |  |
| A1: Ref allele; A2: Effect allele; EAF: Effect allele frequency; HR: Hazard ratio; CI: Confidence Interval; Position: Genomic coordinates. |  |  |  |  |  |  |  |  |  |  |  |  |  |

**Table 2.** Sensitivity analyses in the GEN-SEP study of the three genome-wide significant variants. The models adjusted for the indicated variables plus gender, age and the two main principal components.

|  |  | rs34896991 |  | rs146257041 |  | rs138347802 |  |
| --- | --- | --- | --- | --- | --- | --- | --- |
|  | N | HR (95% CI) | p-value | HR (95% CI) | p-value | HR (95% CI) | p-value |
| BMI | 364 | 6.68 (3.22-13.86) | 3.47x10<sup>-7</sup> | 5.64 (2.10-15.18) | 6.12x10<sup>-4</sup> | 5.72 (0.72-45.32) | 0.098 |
| SAPS | 264 | 4.06 (1.54-10.72) | 4.66x10<sup>-3</sup> | 9.95 (4.55-21.74) | 8.32x10<sup>-9</sup> | 8.63 (1.86-40.01) | 5.90x10<sup>-3</sup> |
| APACHE II | 672 | 3.48 (2.05-5.91) | 3.66x10<sup>-6</sup> | 5.84 (3.43-9.94) | 7.39x10<sup>-11</sup> | 8.65 (4.27-17.53) | 2.07x10<sup>-9</sup> |
| Comorbidities<sup>§</sup> | 600 | 4.24 (2.46-7.33) | 2.19x10<sup>-7</sup> | 5.39 (3.09-9.40) | 2.91x10<sup>-9</sup> | 7.60 (3.14-18.36) | 6.69x10<sup>-6</sup> |
| SOFA | 661 | 4.58 (2.72-7.69) | 9.49x10<sup>-9</sup> | 5.10 (2.99-8.71) | 2.40x10<sup>-9</sup> | 8.50 (4.16-17.39) | 4.59x10<sup>-9</sup> |
| Sepsis of pulmonary origin | 681 | 4.32 (2.62-7.13) | 9.74x10<sup>-9</sup> | 4.71 (2.78-8.00) | 9.33x10<sup>-9</sup> | 8.08 (3.98-16.43) | 7.54x10<sup>-9</sup> |
| Pathogen, % |  |  |  |  |  |  |  |
| Gram-positive | 487 | 4.97 (2.68-9.20) | 3.51x10<sup>-7</sup> | 5.36 (3.02-9.51) | 9.82x10<sup>-9</sup> | 13.11 (5.01-34.32) | 1.61x10<sup>-7</sup> |
| Gram-negative | 487 | 4.97 (2.68-9.21) | 3.42x10<sup>-7</sup> | 5.32 (3.00-90.46) | 1.18x10<sup>-8</sup> | 13.17 (5.04-34.43) | 1.47x10<sup>-7</sup> |
| Others* | 487 | 4.91 (2.64-9.13) | 4.73x10<sup>-7</sup> | 5.13 (2.89-9.12) | 2.30x10<sup>-8</sup> | 12.91 (4.93-33.85) | 1.96x10<sup>-7</sup> |

<sup>§</sup>Includes: cancer, age >80 years old, hepatopathy, valvular disease, immunodeficiency, severe brain damage, morbid obesity, chronic disease, autoimmune disease, pregnancy, myopathy, pneumonia, and serious recurrent infections.
\*Includes: mixed Gram-positive and Gram-negative infection, fungi, virus, and polymicrobial.
APACHE II, Acute Physiology and Chronic Health Evaluation II; BMI, Body Mass Index; ICU, Intensive Care Unit; SAPS, Simplified Acute Physiology Score II; SOFA, Sequential Organ Failure Assessment.

Finally, we tested previously associated variants with sepsis mortality in the results of the first stage. We found that leading variants of previous GWAS^12-14^ were not replicated in the GEN-SEP study (*FER* rs4957796, Hazard Ratio [HR]: 1.09 [95%CI=0.83-1.43], *p*=0.550; *VPS13A* rs117983287, HR: 0.36 [95%CI=0.09-1.48], *p*=0.158; and *CISH* rs143356980, HR: 1.42 [95%CI=0.61-3.33], *p*=0.419) (**Table S6**). Similarly, an assessment of the association results in and around (±50 kb) the corresponding genes (*FER, VPS13A, and CISH*) did not reveal any significant finding (**Supplementary Methods; Table S6**). Likewise, none of classical HLA alleles (*p*_lowest_=0.0169) or amino acids (p_lowest_=0.0169) significantly associated with sepsis survival after multiple testing adjustments (**Figure S5; Table S7**), suggesting that common genetic variation at the HLA is not a major driver of sepsis survival or has a modest effect size that could not be detected with the current design.

### Potential biological effects of the 28-day sepsis survival-associated variants

We then explored the potential functional implications of the three genome-wide significant variants, rs34896991 in *SAMD9*, rs146257041 intergenic to *SLC5A12*\*FIBIN*, and rs138347802 intergenic to LINC00378\MIR3169. Based on distinct functional annotations, we observed a few regulatory activities linked to all three overlapped promotor or enhancer regions in multiple cell-types including blood cells, and T cells (**Table S8**). According to GTEx data, LINC00378 and MIR3169 are expressed only in testis, *SLC5A12* is mainly expressed in the kidney and in the small intestine, *FIBIN* is mainly expressed in arteries (aorta and tibial), tibial nerve, and vagina, while *SAMD9* is expressed broadly across many tissues, but mainly in the esophagus (mucosa), in transformed lymphocytes, and in whole blood.

No significant expression quantitative trait loci (eQTLs) were identified in GTEx for rs34896991 and rs138347802. Nevertheless, four significant eQTLs were found in brain and testis for rs146257041. None of these three variants obtained a significant score predicted using DeepSEA. A scan for previously reported trait associations for the three variants based on PhenoScanner found that rs34896991 in *SAMD9* was also associated with the cause of death in other specified degenerative diseases of the nervous system (**Table S8**). Interestingly, other variants in *SAMD9, SLC5A12, FIBIN*, and in the non-coding RNA (LINC00378) have also been associated with different causes of death according to PhenoScanner results. Regarding the non-coding RNAs, LINC00378 has Cyclin Dependent Kinase Inhibitor 1A (*CDKN1A*) as the main target and is linked to different types of cancers, while MIR3169 targets genes that are mainly involved in the p53 signaling pathway.

To further evaluate the biological implications of the genes near the identified GWAS loci, whole-blood transcriptomic array data from sepsis survivors and non-survivors were assessed. While information was only available for coding genes, an upregulation of *SAMD9* expression in non-surviving sepsis patients was observed (log fold change: 0.545 adjusted FDR *p-*value: 2.18×10^−3^) (**Figure 3**). This gene expression difference among the sepsis patient groups was unrelated to the presence of ARDS (log fold change: 0.011; adjusted FDR *p-*value: 0.996) (**Figure S6**).

**Figure 3.**
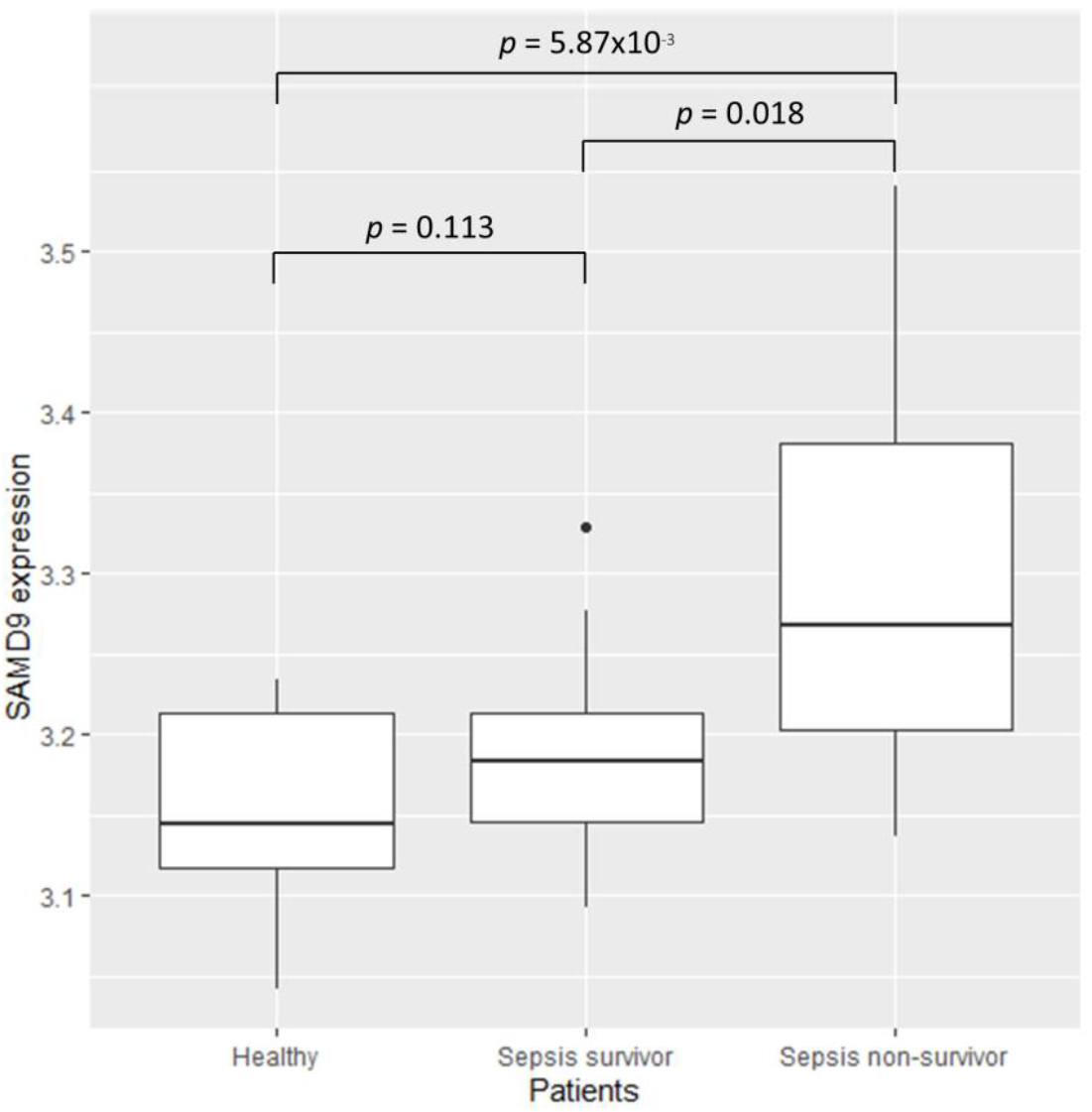
Boxplot of SAMD9 gene expression values among sepsis patients. Significance was calculated by two-sample t-test.

### Polygenic risks of sepsis and effects on 28-day sepsis survival

Finally, we used PRS to investigate whether the polygenic component of sepsis risk was associated with 28-day sepsis survival. We found that the sepsis risk PRS was not significantly associated with sepsis survival at any of the cut-offs (**Figure S7**). These results were similar when the models excluded the variants significantly associated with sepsis in the previous candidate gene studies or in the sepsis mortality GWAS.

## Discussion

To our knowledge, we report the first GWAS of 28-day sepsis survival conducted to date. In addition, given the importance of the MHC in inflammatory and immunological diseases, we also assessed for the first time the association of the classical HLA alleles and amino acids with 28-day sepsis survival. Our findings revealed three novel loci associated with reduced 28-day survival among sepsis patients: one in *SAMD9* (the p.Ala1556Thr exonic variant), one intergenic to *SLC5A12\FIBIN*, and another intergenic to LINC00378\MIR3169. The functional annotation analyses revealed a modest regulatory activity of the sentinel variants. Nevertheless, there was a significant upregulation of *SAMD9* expression in whole blood among non-surviving patients with sepsis in independent whole blood transcriptomic data. We were unable to replicate the findings from previous GWAS of sepsis mortality. We also found a lack of overlap between the polygenic component of sepsis risk and sepsis survival.

*SAMD9* plays a critical role in the inflammatory response during tissue injury and apoptosis^21-22^. This gene encodes one of the SAM domain-containing proteins that has diverse roles for cellular processes via polymerization and participates in protein interactions and RNA binding^23-25^. It has been observed that the *SAMD9* upregulation triggered an accumulation of macrophages increasing low grade glioma progression^26^. Linked to this, it was observed that *SAMD9* interacts with Ral Guanine Nucleotide Dissociation Stimulator Like 2 (RGL2) to decrease the expression of Early growth response protein 1 (EGR1)^27^, which is a key regulator of inflammation in human macrophages^28^. *SAMD9* has been found to be significantly upregulated *in vivo* in peripheral blood mononuclear cells during inflammation and *in vitro* during T cell activation, and its expression is regulated at both the genetic and epigenetic levels^29^. Therefore SAMD9 could serve as a T cell activation marker acting as an anti-inflammatory factor^29^. Other studies support its antitumoral and antiviral activity^22,30- 31^, suggesting that *SAMD9* is part of an evolutionarily conserved defence network hub that has not yet been fully characterized^21^. The osmotic shock and interferon-gamma (IFN-γ) tightly regulate *SAMD9* expression^27,32-33^. In fact, Chefetz and colleagues observed that SAMD9 was up-regulated by tumor necrosis factor-alpha (TNF-α) through p38 mitogen-activated protein kinases (p38 MAPKs) and nuclear factor-kappa-B (NF-κB)^32^. Moreover, mutations in *SAMD9* have been linked to immunodeficiency, neutropenia, impaired anti-cytomegalovirus response, and gastrointestinal disorder^34^, and to severe multisystem disorders and complex phenotypes characterized by recurrent infection, dysphagia, and profound deafness^35^. *SAMD9* is also strongly associated with mean corpuscular hemoglobin or volume and red cell distribution width^36-37^. Therefore, we could hypothesize that the *SAMD9* upregulation, which is inducible by various inflammatory, immunological and stress factors, might activate T cells and produce the accumulation of macrophages through its interaction with RGL2, thus conferring protection against the systemic dysregulation that occurs during sepsis. This would also be supported by its anti-tumoral, anti-inflammatory, and anti-viral activity. Because the T allele of rs34896991 predicts a missense change in *SAMD9*, this allele could act a defective variant explaining its association with increased mortality risk among the patients with sepsis.

Our results also revealed two intergenic variants significantly associated with sepsis survival. One of them was located between *SLC5A12* and *FIBIN* genes. In particular, *SLC5A12* encodes an apical cell membrane protein that acts as an electroneutral and low-affinity sodium (Na(+))-dependent sodium-coupled solute transporter^38^ and it is related to the transport of glucose and other sugars, bile salts and organic acids, metal ions and amino compounds, and the nuclear factor E2-related factor 2 (NRF2) pathway^39-40^. This last pathway is activated under conditions of oxidative stress and has an important role in inflammation^41-43^. FIBIN is a secreted protein essential for pectoral fin bud initiation and T-Box Transcription Factor 5 (tbx5) expression in zebrafish and it has been identified also in mice and humans, acting downstream of retinoic acid and Wnt signalling^44-45^. The other intergenic variant was located between a long non-protein coding RNA (LINC00378) and a microRNA (MIR3169). Although the functional information of these two non-coding genes is scarce, we found that the main LINC00378 target gene is *CDKN1A*, a cell cycle inhibitor involved in terminal differentiation, stem cell renewal, apoptosis, and cell migration that plays a critical role in the cellular response to DNA damage^46^. Different genes are targeted by MIR3169, which are enriched in the p53 signalling pathway, involved in genome stability by preventing mutations caused by cellular stress or DNA damage^47^.

We acknowledge some strengths and limitations of the study. Among the strengths, the results were supported by two independent geographically distinct studies with diverse ancestries. In addition, both cohorts were prospectively enrolled using consensus criteria for sepsis. Because of that, a robust sensitivity analysis was possible to control for potential confounders. Linked to this, although the significance and the effect direction of the associated variants were not affected by the index event bias correction, all the cases used in the survival analysis were also used to assess sepsis risk and this constitutes a limitation for the approach. The main weaknesses of the study are that we could not assess rarer or structural variants in the analyses. Other approaches such as exome or whole-genome sequencing are needed to analyse the role of these rare genetic variants. In addition, further functional characterization of the prioritized variants will be needed to further dissect the mechanistic connections with the pathophysiology of sepsis.

## Conclusion

In conclusion, we have completed a GWAS of 28-day sepsis survival and have identified three novel variants associated with reduced survival, one of them involving a missense variant. These results could potentially allow to identify novel targets for sepsis treatment and patient risk stratification.

## Data Availability

All data produced in the present study are available upon reasonable request to the authors.

## Contributors

TH-B: data analysis, figures and interpretation of data, and drafting the manuscript; BG-G: performed experiments, data analysis and revising the manuscript; JML-S, ES-P, RF, LAR-R, MP, RC-G: statistical analysis and revising the manuscript; ACo, MIG-L, MIP-G, AR-P, DC, JB, AA, EG-H, EE, AM, ET, MMM, LL, DD, AGL, HMG, JPR, TKJ, JMA, and MS: perform experiments, sample and clinical data collection and data analysis; ACa, LVW, JV: interpretation and critical manuscript review; NJM: human subject enrollment, data collection and analysis, cohort funding; interpretation and critical manuscript review; CF: contributed to the study concept and design, data analysis, interpretation of the data, drafting and critical revision of the manuscript, obtained funding, and conceived the project.

## Acknowledgements

We would like to thank all patients for their willingness to participate in the study. We thank all the investigators of the GEN-SEP Network and the nurses who assisted in the collection of donor samples. Similarly, we thank all members of the MESSI study groups for their willingness to support data sharing. TH-B, JML-S, ES-P, LAR-R acknowledge the University of La Laguna for the training support during their PhD studies.

