## Supplemental material for "A Genome-Wide Association Study of Survival in Patients with Sepsis"

**Supplementary methods**

**GEN-SEP study population and samples**

The GEN-SEP study^1^, used for the first stage, is a national, multicenter, observational study conducted in a Spanish network of post-surgical units and ICUs recruiting unrelated adult (>18 years old) patients of European ancestry. This cohort included patients admitted in the following hospitals from Spain: Hospital Universitario de Canarias, Tenerife; Hospital Universitario NS de Candelaria, Tenerife; Hospital Universitario Río Hortega, Valladolid; Hospital Universitario Dr. Negrin, Gran Canaria; Hospital General de Ciudad Real, Ciudad Real; Complejo Hospitalario Universitario de León, León; Hospital Virgen de la Luz, Cuenca; Complejo Hospitalario Universitario de Santiago de Compostela, Santiago de Compostela; Fundació Althaia-Manresa, Barcelona; Hospital Clinic, Barcelona; Hospital Clínico de Valladolid; Hospital La Paz, Madrid; Hospital Fundación Jiménez Díaz, Madrid; Hospital del Bierzo, Ponferrada; Hospital General Río Carrión, Palencia, and Hospital Virgen de la Concha, Zamora. All participants or their representatives gave written informed consent, and the study was approved by the Ethics Committee for Drug Research from the Hospital Universitario de Canarias (Code: CHUNSC_2018-16).

Illustra™ blood genomicPrep Mini Spin Kit was used to purify DNA from peripheral blood for GEN-SEP samples and the concentration was measured on the Qubit 3.0 fluorometer with the dsDNA HS Assay kit (Thermo Fisher Scientific).

**MESSI study population**

Subjects from MESSI study were eligible if they were admitted to the medical ICU with infection-related organ dysfunction consistent with severe sepsis by Sepsis-2 criteria to maintain consistency with cohort initiation^2^. More than 95% of subjects met Sepsis-3 criteria also, but organ dysfunction beyond that captured by the sequential organ failure score was permitted. Subjects were excluded for chronic critical illness based on residence in a long-term acute care hospital, for prior enrollment to the cohort, or if they desired only palliative measures on ICU admission.

**Genotyping and quality control**

Genotyping in the GEN-SEP cohort was performed using the Axiom Genome-Wide Human CEU 1 array (Thermo Fisher Scientific) in the National Genotyping Center (CeGen), Universidad de Santiago de Compostela Node, Spain. The variant calling was achieved following the quality controls recommended by the manufacturer with AffyPipe v2.10.0^3^. Subsequently, the genotyping quality controls were performed with PLINK v1.07^4^ and R v3.6.0 to filter out samples with genotype call rate (CR) <95% and with evidence of relatedness with other individuals (PIHAT>0.2) (**Figure 1**). Single nucleotide polymorphisms (SNPs) from mitochondrial DNA, chromosome Y, or with a genotyping CR <95%, a minor allele frequency (MAF) <0.01, or deviating from Hardy-Weinberg equilibrium (HWE) expectations (*p*<1.0x10^-6^) were excluded from the study (**Figure 1**). PLINK v1.90^5^ was used to calculate the main axis of genetic variation based on 115,611 (1^st^ recruitment period) or 114,420 SNPs (2^nd^ recruitment period) using principal component (PC) analyses (**Figure S1**).

Variant imputation in the GEN-SEP study was performed with the Michigan Imputation Server using the Haplotype Reference Consortium panel (v1.1 2016)^6^. To obtain a better coverage of the Human Leukocyte Antigen (HLA) region, we imputed the classic HLA alleles and amino acids using SHAPEIT v2.837^7-8^ for phasing haplotypes and Impute2 v2.3.2^9^ using the T1DGC HLA Reference Panel with SNP2HLA^10^. Variants with MAF <0.01 or with a low imputation quality (Rsq <0.3) were excluded from the analysis. The X chromosome was analysed separately for men and women, following all quality controls described above, although with SNP filtering based on HWE deviations calculated from females, and then meta-analysed.

In MESSI, SNPs were genotyped using the Axiom TxArray v.1 (Thermo Fisher Scientific). SNPs were filtered out if they were on sex chromosomes, had a MAF <0.01, deviated from HWE (p<1.0x10^-6^), or had a genotyping call rate <95%. Pre-phasing and variant imputation were conducted on Michigan Imputation Server using Minimac3 v1.0.5 or Minimac4 v1.5.7 and The 1000 Genomes Project Phase 1 v3 CEU or CAAPA African American Panel as the reference panel.

**Genome-wide association study of 28-day sepsis survival**

In the first stage, we used the gwasurvivr v1.5.0^11^ package in R to test genetic associations. The meta-analysis of the two GEN-SEP recruitment periods was performed with METASOFT v2.0.1^12^. The independence of prioritized variants was assessed by linkage disequilibrium (LD)-based clumping using PLINK v1.90^5^.

Finally, in the second stage, the meta-analysis from the GEN-SEP and MESSI studies was performed with METASOFT v2.0.1^12^.

**Index event bias assessment**

Since we know that case-only analyses can be biased by variants that are associated with disease risk, we used the Index Event Bias correction^13^. This method uses effect sizes and standard errors for predictors of an index trait and a subsequent trait. This function adjusts the statistics for the subsequent trait for selection bias through the index trait. Thus, we used the results from the sepsis risk study to use the beta and standard error values and apply the method described above. Using a total of 112,363 independent SNPs and the SIMEX method (with 1,000 simulations performed in each stage of the Simex adjustment), we obtained a maximal coefficient of 0.0116 that was used to correct the effect size and the *p-*value of the top hits from our survival GWAS.

**Annotation of the functional effects of associated variants and related genes**

LDproxy module from LDLink v4.1.0 was used to obtain proxies of the sentinel variants in European reference data^14^. Then, we used Genotype-Tissue Expression Project (GTEx) v7 datasets to obtain the local expression quantitative trait loci (eQTLs)^15^. HaploReg v4.1^16^, RegulomeDB v2.0.3^17^, and The Open Targets Post-GWAS portal^18^ were used to annotate their regulatory potential and to rank their functional roles. To quantify the deleterious effects of noncoding variation, we used SNPDelScore^19^. We also analyzed long-distance genomic interactions with Capture Hi-C Plotter (CHiCP)^20^. Moreover, to assess the clinical interpretation of genetic variants we used InterVar, VarSome and ClinVar, as well, LoFtool score to evaluate the gene intolerance based on the ratio of LoF (stop-gain, splice site disrupting, frameshift) mutations to synonymous variants from the Exome Aggregation Consortium (ExAC) dataset. The Variant Effect Predictor (VEP) toolset from Ensembl was used to annotate and predict coding and non-coding regions and we used DeepSEA for a functional significance score prediction. In addition, PhenoScanner v2 was used to facilitate the cross-referencing of genetic variants with a broad range of phenotypes.

Finally, to evaluate the functional and gene targets for the non-coding RNA genes, we assessed the LncRNA2Target database^21^, the LncRNA and Disease Database (version 2.0)^22^, the miRTarBase Release 8.0^23^ and the Kyoto Encyclopedia of Genes and Genomes (KEGG).

**Gene expression of related genes**

To assess differential gene expression of the genes near the sentinel variants, we accessed the public gene expression data GSE54514^24^, available in the Gene Expression Omnibus (GEO) repository. This accession contains transcriptomic information for healthy donors (n=18), and from sepsis survivors (n=26) and non-survivors (n=9). Moreover, we accessed another expression microarray data set (GSE32707)^25^ from sepsis patients (n=30) and patients with ARDS characterization (n=18) to assessed whether ARDS could be an alternative explanation of the gene expression observed differences. A two-sample t-test was used for comparisons, considering a False Discovery Rate (FDR) (q-value) to declare significance while adjusting for comparisons^26^.

**Association of polygenic risks of sepsis with the 28-day sepsis survival**

Polygenic risk scores (PRS) allow for examining the cumulative effect of many genetic variants to be studied. For each individual, the PRS was calculated as the number of risk alleles carried, multiplied by the effect size of the variant, summed across all variants included in the score, i.e.:

$$\mathrm{PRS}_{j}=\sum_{i=1}^{n} \beta_{i}X_{ij}$$

where βi is the log(OR) of variant *i*, *Xij* is the genotype of variant *i* for person *j* and n is the number of variants included in the PRS. We investigated if the polygenic component of sepsis susceptibility was associated with 28-day sepsis survival using PRSice v1.25 software^27^.

Given the lack of GWAS of sepsis risk in the literature, we firstly obtained genome-wide information of sepsis susceptibility comparing the genetic data from the 839 sepsis cases with available information from the GEN-SEP to 1,453 population controls. The controls declared having two generations of ancestors born in Spain and no personal or family history of infectious, cancerous, mental, cardiovascular or immunological diseases. The data from all these controls were used and described in previous studies^28-29^.

For the controls, the array, quality controls, and imputation steps were equivalent to those of the GEN-SEP.

For this GWAS of sepsis susceptibility, association analyses were performed with logistic regression models using EPACTS v3.2.6^30^, adjusting for sex, age, and the two main PCs (not resulting in major systematic deviations from the null; λ=1.04).

For the PRS analyses, a total of 370,316 independent variants were selected after LD-clumping (r^2^≤0.1). We repeated this by excluding all variants within ±500 kb of the candidate genes that were associated with sepsis susceptibility in more than 20 publications (**Table S2**) according to the Public Health Genomics and Precision Health Knowledge Base (v6.2.3) database records. In this case, 368,724 independent variants were used. We also repeated this by excluding all variants within ±500 kb of the sentinel variants (rs4957796, rs117983287, and rs143356980) that were associated with sepsis mortality in previous GWAS^31-33^, leaving a total of 370,063 independent variants for further analysis.

Then, from the different PRS models, we included those variants that met a certain *p-*value threshold and varied this threshold to investigate the effect of including more variants in it. The PRS was tested to identify whether it was associated with 28-day sepsis survival, adjusting for sex, age, and two main PCs. We used Cox regressions with the ‘survival’ package in R v3.6.3 and the previously suggested threshold of *p*<0.001^27^ for defining a significant PRS (**Figure S7**).

**Replication of genes from previous sepsis mortality GWAS**

We evaluated whether the sentinel variants (rs4957796, rs117983287, and rs143356980) previously associated with sepsis mortality in other GWAS^31-33^ were associated with 28-day sepsis survival in the GEN-SEP cohort. Complementarily, we extracted ±50 kb of the regions around the genes where these variants reside (*CISH, FER* and *VPS13A*) and the Genetic Type I error calculator (GEC) was used to estimate the number of independent SNPs for these three regions. This supported the existence of 875 independent variants in Europeans considering the three regions together. Thus, we established the significance threshold at *p*=5.71x10^-5^ for this analysis.

**Supplementary Figures**

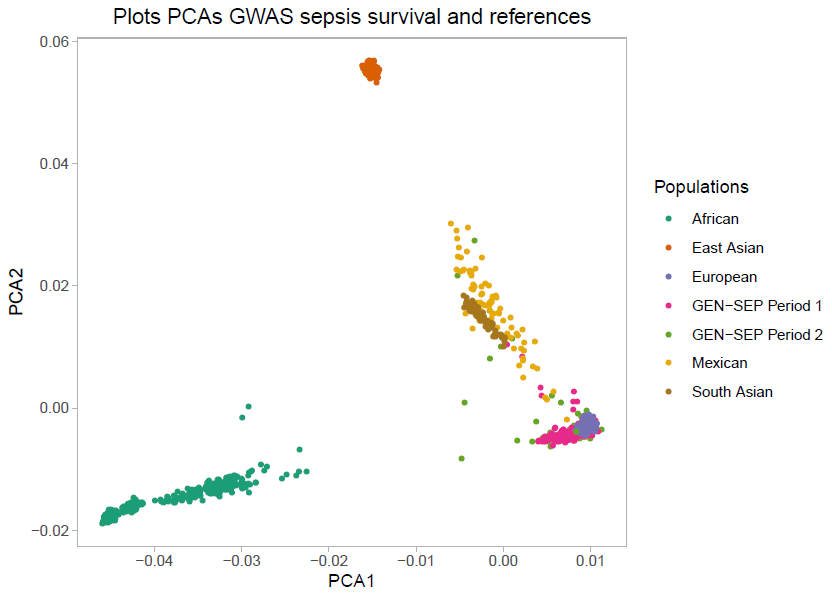

***Supplementary Figure 1.*** *Principal component analysis. Plot of the first two principal components (PCs) of genetic variation of GEN-SEP individuals analysed in the first stage projected on the HapMap3 reference dataset.*

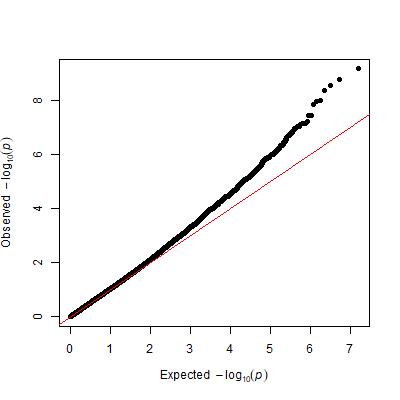

***Supplementary Figure 2.*** *Quantile-quantile plot of the observed versus expected -log10 p-values of the association results of first stage.*

A

B

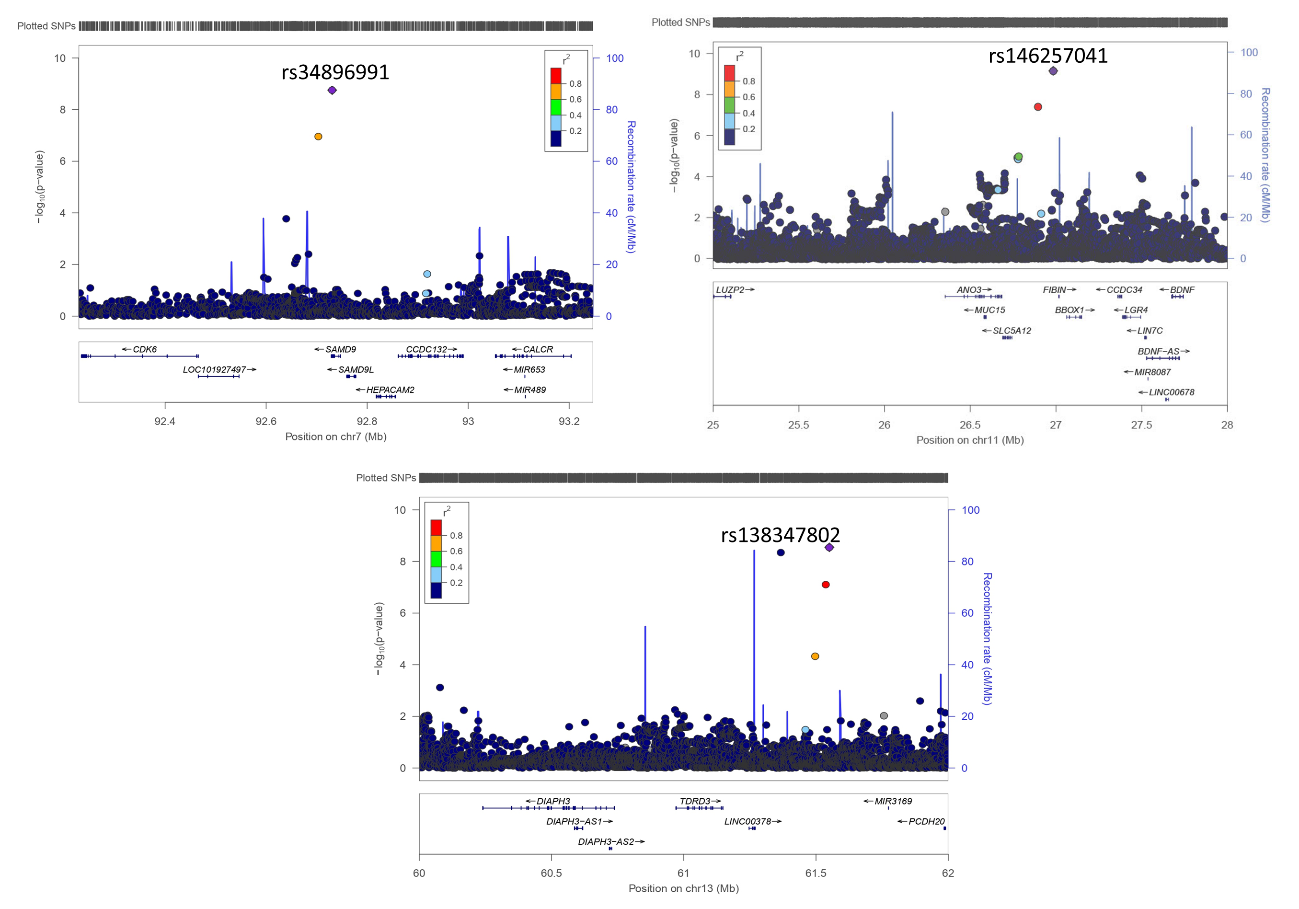

C

***Supplementary Figure 3.*** *Regional plots for the genome-wide significant variants rs34896991 (A), rs146257041 (B), and rs138347802 (C) in the first stage. The x-axis shows the hg19 genomic positions and the left y-axis represents the -log10(p-value). Estimated recombination rates (light blue line) are also plotted on the right y-axis. Linkage disequilibrium values with respect to the sentinel variant (indicated) based on pairwise r2 values in Europeans from the 1000 Genomes Project are shown. Obtained with LocusZoom.*

B

A

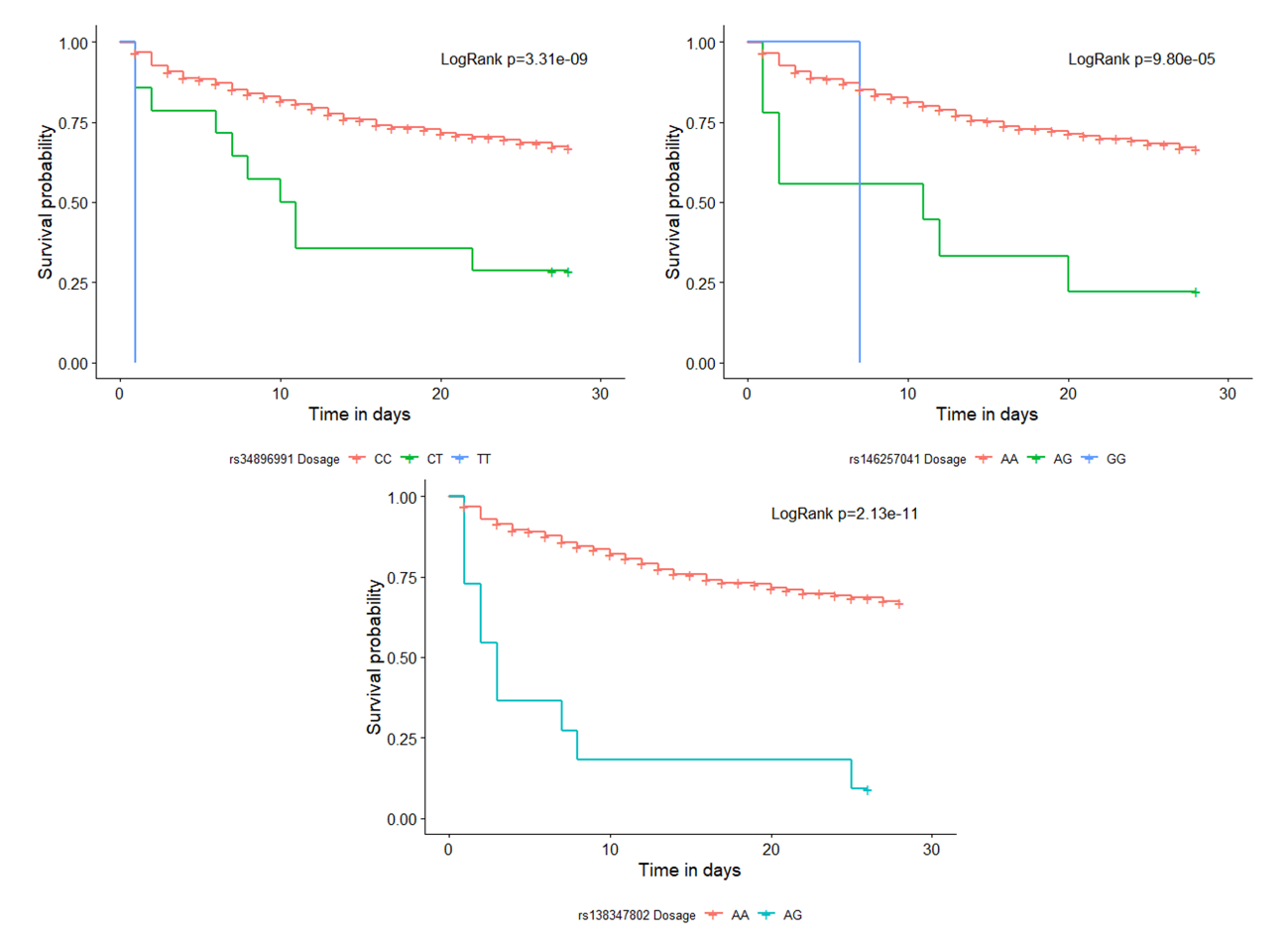
 ***Supplementary Figure 4.*** *Kaplan-Meier 28-day survival plots and the log-rank p-value for the genotypes of the genome-wide significant variants on the first stage: A) rs34896991 (in SAMD9); B) rs146257041 (in SLC5A12\FIBIN); and C) rs138347802 (in LINC00378\MIR3169).*

C

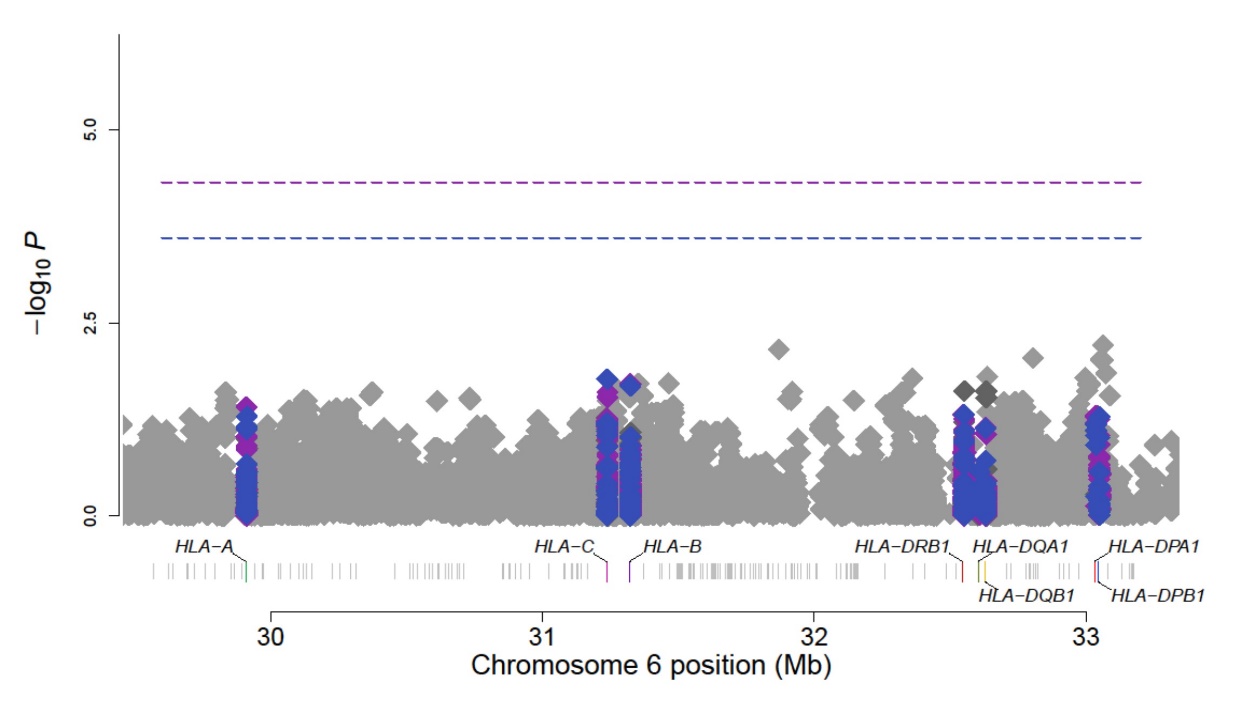

***Supplementary Figure 5.*** *Manhattan plot of sepsis 28-day survival association study results at the HLA region. Highlighted in blue are the classical HLA alleles results, in purple the amino acids results, and in grey the GWAS results of the SNPs (chr6: 28,477,797-33,448,354). Horizontal lines indicate the significance threshold for the different tests: In blue, the threshold for classical HLA alleles p=2.49x10^-4^; and in purple, the threshold for amino acids p=4.83x10^-5^. The x-axis shows genomic position and y-axis represents the significance (-log10[p-value])*. *The association results were represented using the HLA Analysis Tool-Kit (HATK)*^34^*.*

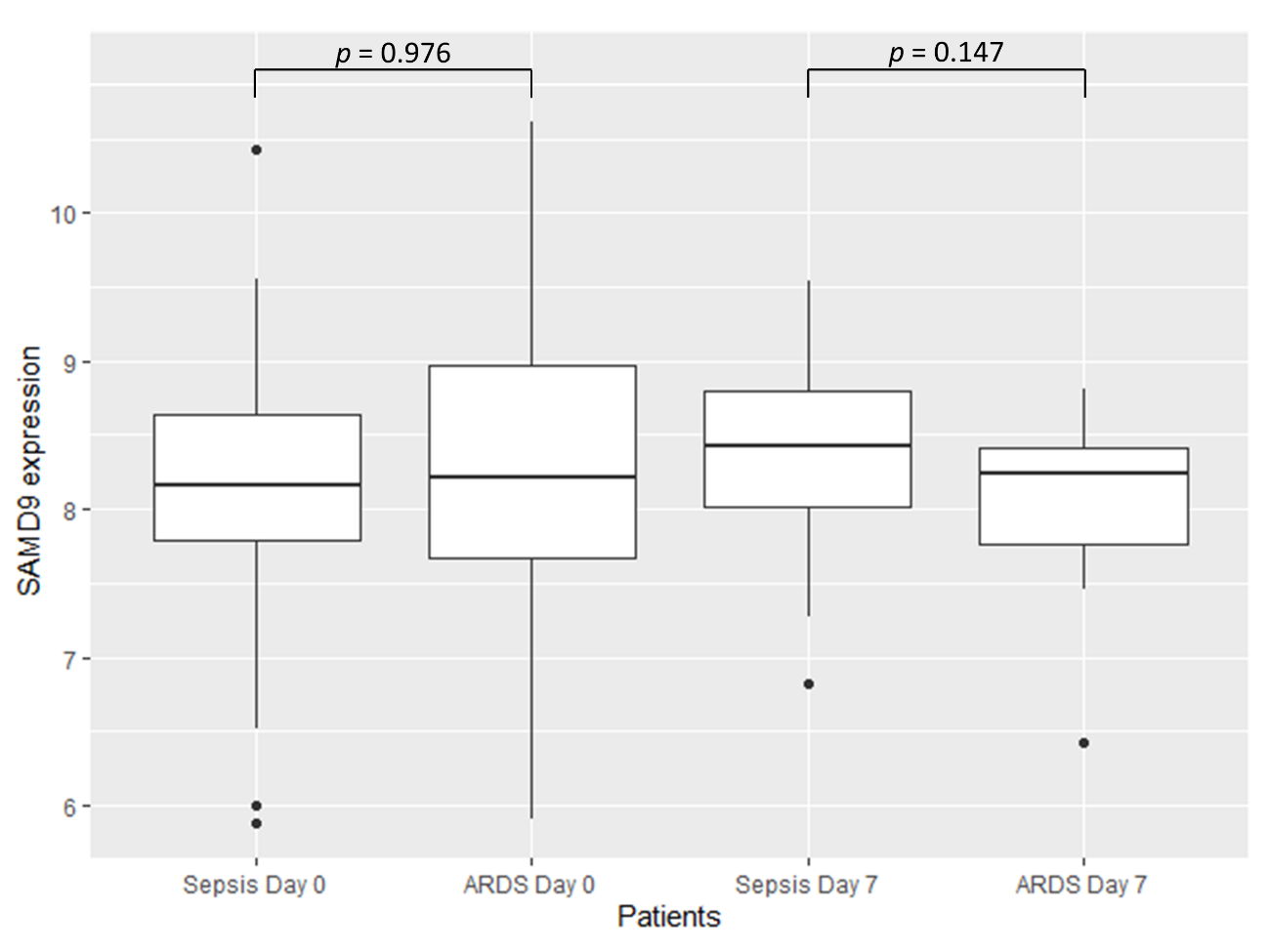

***Supplementary Figure 6.*** *Boxplot of SAMD9 gene expression values in GSE32707.* *Significance was calculated by two-sample t-test.*

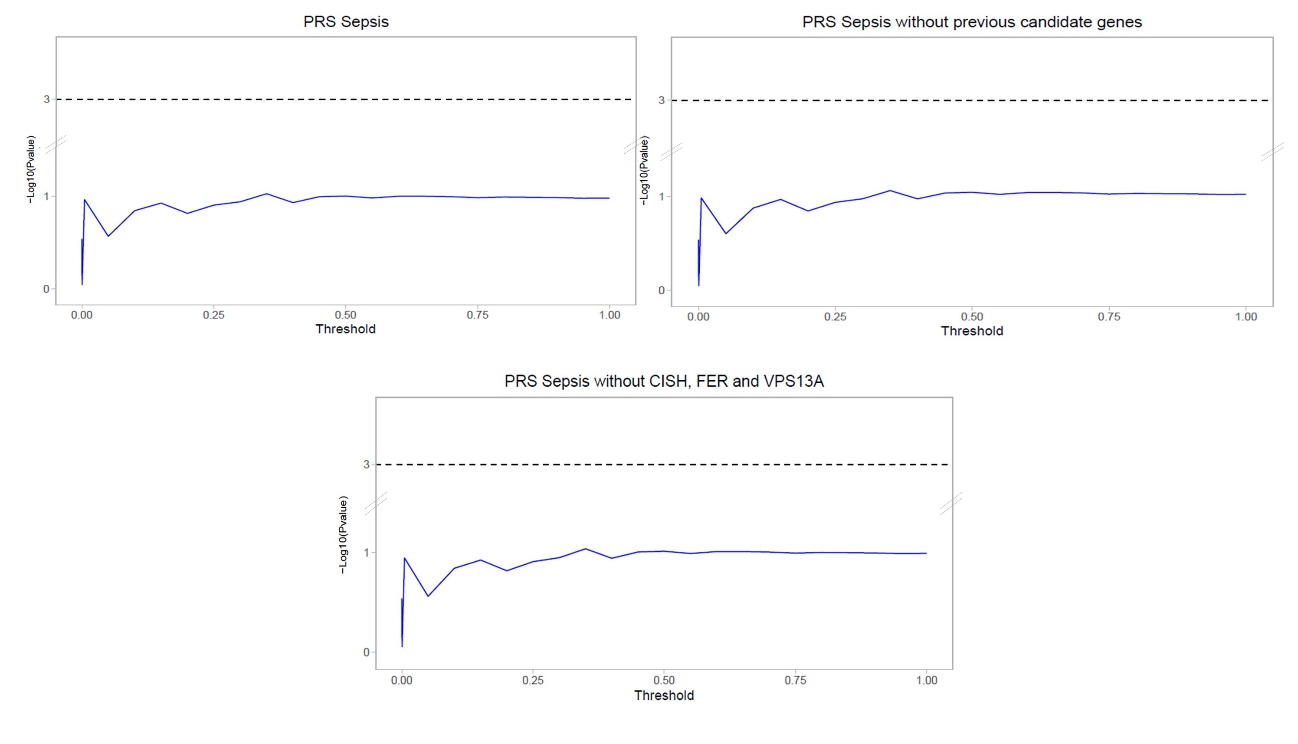

C

B
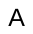

A

***Supplementary Figure 7.*** *Polygenic risk score (PRS) model fitting and strength of association in the GEN-SEP patients with available mortality data at 28 days after sepsis diagnosis using the Cox regression p-values. The x-axis shows the threshold p-value used for selecting the variants to be included in the risk score model. The y-axis shows the significance (−log10 (p-value)) for the association of the risk score models with 28-day survival for each threshold used. Horizontal lines indicate the significance threshold. A) All-inclusive PRS model fitting. B) PRS models fitting excluding variants (+/- 500 kb) linked to sepsis risk based on candidate gene studies (see* ***Table S2****). C) PRS model fitting excluding the variants (and +/- 500 kb proxies) of sepsis mortality genes (FER, VPS13A, and CISH) described in previous GWAS.*

**Supplementary Tables**

| **Table S1. Relevant demographic and clinical features of cases and controls used for the GWAS of sepsis risk after quality controls.** | | | |
| --- | --- | --- | --- |
|  | **Cases (N = 832)** | **Controls (N = 1,443)** | ***p*-value*** |
| Sex, % male (N) | 63 (524) | 49 (703) | < 0.0001 |
| Age, mean years ± SD | 63 ± 15 | 47 ± 14 | < 0.0001 |
| BMI, mean kg/m2 ± SD | 28 ± 6 | NA | NA |
| SAPS, mean ± SD | 47 ± 15 | NA | NA |
| APACHE II (8h), mean ± SD | 20 ± 7 | NA | NA |
| SOFA, mean ± SD | 8 ± 4 | NA | NA |
| Comorbidities^$^, % (N) | 46 (326) | NA | NA |
| ARDS, % (N) | 38 (318) | NA | NA |
| Hospital mortality, % (N) | 35 (287) | NA | NA |
| ICU mortality, % (N) | 28 (228) | NA | NA |
| Days in Hospital, mean ± SD | 38 ± 50 | NA | NA |
| Days in ICU, mean ± SD | 17 ± 24 | NA | NA |
| Sepsis of pulmonary origin, % (N) | 35 (264) | NA | NA |
| Pathogen, % (N) |  |  |  |
| Gram-positive | 26 (150) | NA | NA |
| Gram-negative | 37 (209) | NA | NA |
| Others^+^ | 28 (158) | NA | NA |
| *Sex comparison was assessed by a chi-square test. Age was compared using the Mann–Whitney U-test.  ^$^Comorbidities include cancer, age >80 years, hepatopathy, valvular disease, immunodeficiency, severe brain damage, morbid obesity, chronic disease, autoimmune disease, pregnancy, myopathy, pneumonia, and serious recurrent infections.  ^+^Others include both (Gram-positive and Gram-negative), fungi, virus and polymicrobial. APACHE II, Acute Physiology and Chronic Health Evaluation II; ARDS, Acute Respiratory Distress Syndrome; BMI, Body Mass Index; ICU, Intensive Care Unit; SAPS, Simplified Acute Physiology Score II; SOFA, Sequential Organ Failure Assessment. | | | |

| **Table S2.** **Previous candidate genes associated with sepsis risk obtained from the Public Health Genomics and Precision Health Knowledge Base (v6.2.3) database.** | | | | |
| --- | --- | --- | --- | --- |
| **Associated Gene** | **Total Publications** | **Gene name** | **Genomic locations (GRCh37/hg19)** | **Position +/- 500 kb** |
| *TNF* | 102 | Tumour Necrosis Factor | chr6:31,543,344-31,546,113 | chr6:31,043,344-32046113 |
| *IL6* | 65 | Interleukin 6 | chr7:22,765,503-22,771,621 | chr7:22,265,503-23,271,621 |
| *IL10* | 60 | Interleukin 10 | chr1:206,940,947-206,945,839 | chr1:206,440,947-210,445,839 |
| *TLR4* | 53 | Toll Like Receptor 4 | chr9:120,466,460-120,479,768 | chr9:119,966,460-120,979,768 |
| *CD14* | 42 | Monocyte Differentiation Antigen CD14 | chr5:140,011,313-140,013,286 | chr5:139,511,313-140,513,286 |
| *MBL2* | 41 | Mannose Binding Lectin 2 | chr10:54,525,140-54,531,460 | chr10:54,025,140-55,031,460 |
| *IL1B* | 40 | Interleukin 1 Beta | chr2:113,587,328-113,594,480 | chr2:113,087,328-114,094,480 |
| *TLR2* | 34 | Toll Like Receptor 2 | chr4:154,605,441-154,627,243 | chr4:154,105,441-155,127,243 |
| *LTA* | 33 | Lymphotoxin Alpha | chr6:31,539,831-31,542,101 | chr6:31,039,831-32,042,101 |
| *SERPINE1* | 20 | Serpin Peptidase Inhibitor, Clade E (Nexin, Plasminogen Activator Inhibitor Type 1) | chr7:100,770,370-100,782,547 | chr7:100,270,370-101,282,547 |
| *IL1RN* | 22 | Interleukin 1 Receptor Antagonist | chr2:113,864,791-113,891,593 | chr2:113,364,791-114,391,593 |

| **Table S3. Demographic and clinical features of GEN-SEP patients of the first stage GWAS of 28-day survival.** | | | | | | |
| --- | --- | --- | --- | --- | --- | --- |
|  | 1st GEN-SEP period | | | 2nd GEN-SEP period | | |
|  | **Survivors (N=329)** | **Deaths (N=146)** | ***p*-value*** | **Survivors (N=177)** | **Deaths (N=35)** | ***p*-value*** |
| Sex, % male (N) | 64 (212) | 68 (100) | 0.451 | 56 (100) | 51 (18) | 0.715 |
| Age, mean years ± SD | 62 ± 15 | 66 ± 13 | 4.93x10^-3^ | 64 ± 15 | 73 ± 13 | 2.36x10^-3^ |
| BMI, mean ± SD | 28 ± 6 | 26 ± 5 | 0.089 | 27 ± 6 | 26 ± 5 | 0.509 |
| LISS, Mean ± SD | 3 ± 2 | 4 ± 3 | 0.263 | 1 ± 2 | 1 ± 1 | 0.148 |
| SAPS, mean ± SD | 45 ± 13 | 53 ± 16 | 4.56x10^-5^ | 50 ± 16 | 48 ± 26 | 0.892 |
| APACHE II, mean ± SD | 20 ± 7 | 25 ± 7 | 1.72x10^-10^ | 17 ± 7 | 22 ± 7 | 2.64x10^-4^ |
| SOFA, mean ± SD | 8 ± 4 | 9 ± 5 | 3.71x10^-3^ | 7 ± 3 | 10 ± 4 | 7.21x10^-4^ |
| ARDS, % (N) | 44 (145) | 60 (87) | 2.51x10^-3^ | 13 (23) | 26 (9) | 0.096 |
| Comorbidities^$^, % (N) | 41 (118) | 52 (56) | 0.063 | 39 (66) | 46 (16) | 0.606 |
| Sepsis of pulmonary origin, % (N) | 39 (125) | 39 (57) | 0.962 | 23 (40) | 31 (11) | 0.368 |
| Days in Hospital, mean ± SD | 45 ± 40 | 17 ± 12 | 2.23x10^-17^ | 36 ± 49 | 16 ± 13 | 7.20x10^-5^ |
| Days in ICU, mean ± SD | 22 ± 23 | 10 ± 7 | 1.22x10^-7^ | 13 ± 30 | 9 ± 7 | 0.742 |
| Pathogen, % (N) |  |  |  |  |  |  |
| Gram-positive | 30 (59) | 32 (26) | 0.794 | 21 (36) | 15 (5) | 0.561 |
| Gram-negative | 41 (81) | 35 (28) | 0.412 | 30 (52) | 35 (12) | 0.688 |
| Others+ | 30 (59) | 33 (27) | 0.341 | 28 (49) | 38 (13) | 0.343 |
| *Comparisons for sex, ARDS, comorbidities, sepsis of pulmonary origin, and pathogen were conducted by chi-square test. The rest of variables were compared using a Mann–Whitney U test.  ^$^Includes: cancer, age >80 years, hepatopathy, valvular disease, immunodeficiency, severe brain damage, morbid obesity, chronic disease, autoimmune disease, pregnancy, myopathy, pneumonia, and serious recurrent infections.  ^+^Includes: mixed Gram-positive and Gram-negative infection, fungi, virus, and polymicrobial.  APACHE II, Acute Physiology and Chronic Health Evaluation II; ARDS, acute respiratory distress syndrome; BMI, Body Mass Index; ICU, Intensive Care Unit; SAPS, Simplified Acute Physiology Score II; SOFA, Sequential Organ Failure Assessment. | | | | | | |

| **Table S4. Prioritized independent SNPs from the first stage of the GWAS of 28-day sepsis survival.** | | | | | | | | | | | | | | |
| --- | --- | --- | --- | --- | --- | --- | --- | --- | --- | --- | --- | --- | --- | --- |
|  |  |  |  |  |  | GEN-SEP (N=687) |  | MESSI (N=1,362)  (European Americans) | | MESSI (N=701)  (African Americans) | | Meta-analysis (N=2,750) | | |
| **rsID** | **Position (Hg19)** | **A1/A2** | **Function** | **Gene** | **EAF** | **HR (95% CI)** | ***p*-value** | **HR (95% CI)** | ***p*-value** | **HR (95% CI)** | ***p*-value** | | **HR (95% CI)** | ***p*-value** |
| rs114581095 | 3:36047297 | C/T | Intergenic | *ARPP21\STAC* | 0.016 | 6.07 (3.15-11.71) | 7.59x10^-8^ | 1.14 (0.91-1.43) | 0.380 | 0.79 (0.39-1.58) | 0.712 | | 1.79 (1.38-6.35) | 7.84x10^-6^ |
| rs114658749 | 3:195775144 | C/T | Intergenic | *SDHAP1\TFRC* | 0.062 | 3.05 (2.03-4.60) | 9.16x10^-8^ | 0.83 (0.55-1.26) | 0.255 | 1.17 (0.51-2.64) | 0.501 | | 1.44 (1.22-3.02) | 5.56x10^-6^ |
| rs76805442 | 5:84399852 | G/A | Intergenic | *EDIL3\NBPF22P* | 0.066 | 2.41 (1.71-3.39) | 4.37 x10^-7^ | 1.05 (0.83-1.32) | 0.694 | 1.88 (0.99-3.58) | 0.054 | | 1.65 (1.33-3.05) | 3.39x10^-6^ |
| rs73285901 | 7:25189764 | G/A | Intronic | *C7orf31* | 0.032 | 3.19 (2.03-5.01) | 4.76x10^-7^ | 0.99 (0.74-1.34) | 0.968 | 0.89 (0.74-1.07) | 0.199 | | 1.37 (1.37-2.55) | 8.77x10^-5^ |
| rs34896991 | 7:92730745 | C/T | Exonic | *SAMD9* | 0.018 | 4.75 (2.86-7.89) | 1.77x10^-9^ | 0.61 (0.29-1.28) | 0.188 | 1.44 (0.46-4.50) | 0.534 | | 1.64 (1.37-6.78) | 4.92x10^-8^ |
| rs146257041 | 11:26983813 | A/G | Intergenic | *SLC5A12\FIBIN* | 0.015 | 5.14 (3.06-8.65) | 7.04x10^-10^ | NA | NA | 0.89 (0.12-6.39) | 0.911 | | 4.59 (2.77-7.59) | 3.00x10^-9^* |
| rs138347802 | 13:61367197 | A/G | Intergenic | *LINC00378\MIR3169* | 0.014 | 8.33 (4.1-16.91) | 4.56x10^-9^ | 0.68 (0.3-1.52) | 0.345 | 2.98 (0.95-9.38) | 0.062 | | 2.57 (1.82-13.03) | 4.44x10^-8^ |
| rs113925942 | 13:93552176 | T/C | Intergenic | *GPC5\LINC00363* | 0.022 | 4.10 (2.44-6.88) | 9.63x10^-8^ | 1.10 (0.49-2.46) | 0.813 | NA | NA | | 2.21 (1.59-3.07) | 1.34x10^-6^* |
| rs183364907 | 14:58109460 | A/T | Intronic | *SLC35F4* | 0.011 | 7.40 (3.55-15.45) | 9.79x10^-8^ | 0.54 (0.22-1.3) | 0.171 | 2.88x10^-7^ (0-Inf) | 0.992 | | 2.02 (1.51-2.72) | 1.41x10^-6^ |
| rs146077854 | 17:14555200 | G/A | Intergenic | *HS3ST3B1\LOC101928475* | 0.010 | 7.25 (3.36-15.67) | 4.65x10^-7^ | NA | NA | NA | NA | | NA | NA |
| rs1573332 | 21:39902441 | T/A | Intronic | *ERG* | 0.345 | 1.89 (1.48-2.40) | 2.41x10^-7^ | 0.96 (0.85-1.08) | 0.485 | NA | NA | | 1.33 (1.17-1.53) | 1.14x10^-5^* |
| *Meta-analysis performed between two studies.  A1: Ref allele; A2: Effect allele; EAF: Effect allele frequency; HR: Hazard ratio; CI: Confidence Interval; Position: chromosome and base pair. | | | | | | | | | | | | | | |

| **Table S5.** **Results for the 28-day sepsis survival association of the significant SNPs after correcting for the index bias event.** | | | | | | | | | | | | |
| --- | --- | --- | --- | --- | --- | --- | --- | --- | --- | --- | --- | --- |
|  |  |  | **28-day survival model** | | | **Sepsis risk model** | | | **28-day survival with index event bias adjustment** | | | |
| **Gene** | **rsID** | **A1/A2** | **Beta** | **SE** | ***p*-value** | **Beta** | **SE** | ***p*-value** | **Beta** | **SE** | **HR (95% CI)** | ***p*-value** |
| *SAMD9* | rs34896991 | C/T | 1.558 | 0.259 | 1.77x10^-9^ | 0.389 | 0.320 | 0.223 | 1.554 | 0.259 | 4.73 (2.85-7.86) | 9.99x10^-10^ |
| *SLC5A12\FIBIN* | rs146257041 | A/G | 1.638 | 0.266 | 7.04x10^-10^ | 0.675 | 0.355 | 0.057 | 1.630 | 0.266 | 5.10 (3.03-8.59) | 4.39x10^-10^ |
| *LINC00378\MIR3169* | rs138347802 | A/G | 2.120 | 0.362 | 4.56x10^-9^ | 0.202 | 0.405 | 0.617 | 2.117 | 0.362 | 8.31 (4.09-16.88) | 2.40x10^-9^ |
| A1: Ref allele; A2: Effect allele; EAF: Effect allele frequency; HR: Hazard ratio. | | | | | | | | | | | | |

| **Table S6. Association results of the sentinel variants from other sepsis mortality GWAS studies in GEN-SEP and the variant with the lowest *p-*value in the gene region in this study.** | | | | | | | | |
| --- | --- | --- | --- | --- | --- | --- | --- | --- |
| **rsID** | **Study** | **Position (hg19)** | **A1/A2** | **Function** | **Gene** | **EAF** | **HR (95% CI)** | ***p*-value** |
| rs143356980 | Rosier et al. | 3:50638812 | C/T | Intergenic | *HEMK1\CISH* | 0.013 | 1.42 (0.61-3.33) | 0.419 |
| rs78292306 | GEN-SEP | 3:50646863 | G/A | Intronic | *CISH* | 0.021 | 0.57 (0.23-1.41) | 0.224 |
| rs4957796 | Rautanen et al. | 5:108402140 | T/C | Intronic | *FER* | 0.161 | 1.09 (0.83-1.43) | 0.550 |
| rs115761658 | GEN-SEP | 5:108397843 | G/A | Intronic |  | 0.020 | 3.58 (1.83-7.03) | 2.05x10^-4^ |
| rs117983287 | Scherag et al. | 9:80020874 | C/A | Exonic | *VPS13A* | 0.019 | 0.36 (0.09-1.48) | 0.158 |
| rs4745630 | GEN-SEP | 9:80023542 | G/A | Intronic |  | 0.286 | 1.39 (1.12-1.73) | 2.99x10^-3^ |
| A1: Ref allele; A2: Effect allele; EAF: Effect allele frequency from Stage 1; HR: Hazard ratio. | | | | | | | | |

| **Table S7. Nominally significant (*p*<0.05) results for the classical HLA alleles and amino acids in GEN-SEP.** | | | | | | |
| --- | --- | --- | --- | --- | --- | --- |
|  | **Gene** | **Allele/Variant*** | **Genomic position (hg19)** | **Freq.** | ***p*-value** | **HR (95% CI)** |
| Classical HLA alleles | *HLA-C* | 03:03 | 6:31238192 | 0.965 | 0.017 | 0.57 (0.36-0.90) |
|  | *HLA-B* | 15:01 | 6:31323293 | 0.970 | 0.021 | 0.55 (0.33-0.91) |
|  | *HLA-DRB1* | 10 | 6:32552064 | 0.982 | 0.049 | 0.54 (0.29-1.00) |
|  | *HLA-DRB1* | 10:01 | 6:32552064 | 0.982 | 0.049 | 0.54 (0.29-1.00) |
|  | *HLA-C* | Arg91Gly | 6:31347104 | 0.965 | 0.017 | 0.57 (0.36-0.90) |
| Amino acids | *HLA-B* | Trp156 | 6:31432003 | 0.968 | 0.019 | 0.55 (0.33-0.91) |
|  | *HLA-B* | --156 | 6:31432003 | 0.968 | 0.019 | 0.55 (0.33-0.91) |
|  | *HLA-C* | Leu/Trp156 | 6:31346909 | 0.419 | 0.025 | 0.79 (0.64-0.97) |
|  | *HLA-C* | Arg/Gln156 | 6:31346909 | 0.590 | 0.029 | 1.27 (1.02-1.57) |
|  | *HLA-A* | Arg/Trp156 | 6:30019219 | 0.756 | 0.039 | 0.77 (0.61-0.99) |
|  | *HLA-DRB1* | Pro231 | 6:32656010 | 0.982 | 0.049 | 0.54 (0.29-1.00) |
|  | *HLA-DRB1* | Gln166 | 6:32657380 | 0.982 | 0.049 | 0.54 (0.29-1.00) |
|  | *HLA-DRB1* | Arg30 | 6:32660058 | 0.982 | 0.049 | 0.54 (0.29-1.00) |
|  | *HLA-DRB1* | Val31 | 6:32660055 | 0.982 | 0.049 | 0.54 (0.29-1.00) |
|  | *HLA-DRB1* | Ala38 | 6:32660034 | 0.982 | 0.049 | 0.54 (0.29-1.00) |
|  | *HLA-DRB1* | Tyr40Phe | 6:32660028 | 0.982 | 0.049 | 0.54 (0.29-1.00) |
|  | *HLA-DRB1* | Glu10 | 6:32660118 | 0.982 | 0.049 | 0.54 (0.29-1.00) |
| *Amino acid and position; Freq: Frequency of classical HLA allele or amino acids; HR: Hazard ratio. | | | | | | |

| **Table S8. Functional assessment of variants associated with 28-day sepsis survival after meta-analysis.** | | | |
| --- | --- | --- | --- |
|  | **rs34896991** (*SAMD9)* | **rs146257041** *(SLC5A12\FIBIN)* | **rs138347802** (LINC00378\MIR3169) |
| **Chromosome location** | chr7:92730745 (p.Ala1556Thr) | chr11:26983813 (intergenic) | chr13:61367197 (intergenic) |
| **Functional significance score** [DeepSEA *p* ≤ 0.05] | 5.87x10^-2^ | 0.235 | 0.138 |
| **Clinical interpretation of genetic variants** | Likely benign (InterVar); Benign (Varsome and ClinVar) | NA | NA |
| **LoFtool score < 0.1** | 0.991 | None | None |
| **CADD (scaled)** | 10.8 | 1.61 | 6.73 |
| **Variant Effect Predictor (**[VEP]**)** | 1 feature: ENSG00000205413 (missense variant) | None | None |
| **RegulomedB rank (Score)** | Motif hit (6, 0.437) | Other (7, 0.184) | Other (7, 0.184) |
| **Enhancer histone marks** [HaploReg] | None | H3K4me1 (Mesench and Epithelial) and H3K27ac (Blood & Primary T helper memory cells from peripheral blood; Epithelial, Foreskin Keratinocyte Primary Cells skin and Muscle Colon Smooth Muscle). | None |
| **Promoter histone marks** [HaploReg] | H3K4me3 (Blood and T-cell; Mesench; Brain; Adipose; Heart; Muscle; Digestive; Fetal Lung and Liver) | None | H3K9ac (Breast Myoepithelial Primary Cells) |
| **DNAse** [HaploReg] | None | None | None |
| **Altered regulatory motifs** [HaploReg] | Egr-1,Pax-4 | ERalpha-a | 5 altered motifs (AP-1_disc6,  DMRT2, Hoxa7_1, Nanog_disc4 and Pou2f2) |
| **Proteins bound** [HaploReg] | None | None | None |
| **CHICP** | None | NA | NA |
| **Open Targets Genetics**  Top ranked genes based on the overall V2G score | 0.27 (*SAMD9*) | 0.14 (*SLC5A12*) | 0.07 (LINC01442) |
| **dsQTL** [GTEX]  Tissue-specific *p* ≤ 0.05 | None | None | None |
| **eQTL** [GTEX]  Tissue-specific *p* ≤ 0.05 | Not significant | 4 significant eQTLs were found for this variant in brain and testis | Not significant |
| **Score CAPE dsQTL >0.5** | None | None | None |
| **Score CAPE eQTL >0.5** | None | None | None |
| **PhenoScanner**  *p* ≤ 0.05 | Allele T has been related with the cause of death of other specified degenerative diseases of the nervous system (*p*=8.62x10^-11^). | NA | NA |
| CADD, Combined annotation dependent depletion; CAPE, Cellular dependent deactivating; CHiCP, capture HiC plotter; dsQTL, DNase I sensitivity quantitative trait loci; EGR-1, Early growth response protein 1; eQTL, expression quantitative trait loci; PAX-4, Paired box 4; and V2G, Assigning variants to genes. | | | |
